# Does the morphology of the posterior malleolar fragment influence clinical outcomes of trimalleolar fractures after surgical fixation? A systematic review and Meta-analysis

**DOI:** 10.1101/2022.01.05.21268417

**Authors:** Sandeep Patel, Mandeep Singh Dhillon, Vishnu Baburaj, Siddhartha Sharma

## Abstract

**Background:** Posterior malleolus (PM) fractures have historically been classified according to the size of the fragment, to study the need for surgical fixation and to assess clinical outcomes. Recent research has suggested that the morphology of the PM fragment is of more relevance than its size.

**Objectives:** This systematic review aims to determine if the size of the PM fragment influences clinical outcomes of trimalleolar fractures after surgical fixation, and if so, to find out which fracture type has the best outcomes.

**Methods:** This systematic review will be conducted according to the PRISMA guidelines. A literature search will be conducted on the electronic databases of PubMed, Embase, Scopus and Ovid with a pre-determined search strategy. A manual bibliography search of included studies will also be done. Original articles in English that have relevant data on the outcomes of PM fractures and its morphology will be included. Data will be extracted from included studies and analysis carried out with the help of appropriate software.

## 1. Background

Trimalleolar fractures comprise of fracture of medial and lateral malleoli, as well as the posterior aspect of the tibial plafond (posterior malleolus, PM) and account for about 7% of all ankle fractures [1]. The PM fragment results from bony avulsion of the posterior inferior tibiofibular ligament (PITFL) and its presence leads to inferior outcomes in ankle fractures [2, 3].

Haraguchi et al. challenged the historical concept that PM fractures must be grouped according to the size of the fragment to determine the need for surgical fixation and postoperative outcomes [4]. Current literature lacks sufficient information whether the PM fracture morphology actually influences the long-term clinical outcomes of ankle fractures with PM fragment.

## 2. Need for review

The morphology of the PM fragment is shown to be more important than its size in determining need for surgical fixation and outcomes [5]. To the best of our knowledge, there are currently no systematic reviews that compare clinical outcomes between the morphological PM fragment types as classified by Haraguchi et al. In this review, we aim to determine if the PM fracture morphology has any bearing on the long-term functional outcomes, and if so, which fracture type has the best results.

## 3. Objective

To determine if the PM fracture morphology has any bearing on the long-term functional outcomes, and if so, which fracture type has the best results.

## 4. PICO framework for the study

a. Participants : Adult human subjects with ankle fracture involving the posterior malleolus
b. Intervention : Surgical fixation of fracture
c. Control : Not applicable
d. Outcome : Complications, patient-reported outcome scores, ankle arthritis, need for revision

## 5. Methods

This systematic review and meta-analysis will be done according to the Preferred Reporting Items for Systematic Reviews and Meta-analysis (PRISMA) guidelines.

a. Review Protocol A protocol of the review will be prepared as per the PRISMA-P guidelines.
b. Eligibility Criteria Original research on human adult subjects having PM ankle fractures undergoing surgical fixation will be included. The studies must mention the morphology of the PM fragment and clinical outcome parameters. Studies in languages other than English, lower evidence studies such as case reports, case series, animal, cadaveric and biomechanical studies will be excluded.
c. Information Sources and Literature search Electronic databases of PubMed, Embase, Scopus, and Ovid will be searched using the keywords “(Posterior malleolus) OR trimalleolar OR (ankle fracture) OR (unstable ankle) AND (outcome OR Result)” for studies in English published from inception to date of search. A bibliography search of included studies will also be carried out for more potentially eligible articles.
d. Study Selection Two authors will separately go through the title and abstract of the search results to narrow down studies using the inclusion and exclusion criteria. In case of any doubt, the full text of the study will be obtained and assessment made after discussion with all the authors.
e. Data Collection and Data Items Data from eligible studies will be extracted on excel spreadsheets, which will be cross-checked for accuracy. The following data will be collected:

- Name of first author and publication year
- Study design
- Haraguchi fracture classification
- Number of participants and their demographic data
- Outcome scores
- Rate of complications and revision surgeries
f. Outcome measures The outcome measures that would be considered for analysis are as follows:

- Visual analog scale (VAS
- American Orthopaedic Foot and Ankle Society (AOFAS) Score
- Olerud-Molander Ankle Score (OMAS)
g. Data Analysis and Synthesis Both qualitative and quantitative synthesis will be performed, if adequate data is obtained. Meta-analysis would be conducted to compare the pooled estimate of outcomes between various Haraguchi fracture types if reported by more than three studies. A meta-regression of outcomes will also be carried out keeping the Haraguchi subtype as covariates. RevMan version 5.4 (computer program) and OpenMeta Analyst will be used for analysis. A fixed or random-effects model will be chosen based on the amount of heterogeneity. 95% confidence intervals will be used, and results would be depicted using forest plots.
h. Assessment of Risk of Bias The methodological index for non-randomized studies (MINORS) tool will be used to assess bias in observational studies, and RoB 2.0 tool will be utilized for randomized controlled trials [6, 7].

## Data Availability

All data produced in the present work are contained in the manuscript

